# The interplay between air pollution, built environment, and physical activity: perceptions of children and youth in rural and urban India

**DOI:** 10.1101/2023.08.22.23294434

**Authors:** Jamin Patel, Tarun Reddy Katapally, Anuradha Khadilkar, Jasmin Bhawra

## Abstract

The role of physical inactivity as a contributor to non-communicable diseases (NCDs) risk in children and youth is widely recognized. Air pollution and built environment can limit participation in physical activity and exacerbate NCD risk; however, the relationships between perceptions of air pollution, built environment, and health behaviors are not fully understood, particularly among children and youth in low and middle-income countries. Currently, there are no studies capturing how child and youth perceptions of air pollution and built environment influence physical activity in India, thus, this study investigates the association between perceived air pollution and built environmental factors on moderate-to-vigorous physical activity (MVPA) levels of children and youth in both rural and urban India. Online surveys captured MVPA, perception of air pollution and built environment factors, as well as relevant sociodemographic characteristics from parents and children aged 5 to 17 years in partnership with 41 schools across 28 urban and rural locations during the Coronavirus disease lockdowns in 2021. After adjusting for age, gender, and location, a significant association was found between the perception of air pollution and MVPA levels (β = −18.365, p < 0.001). Similarly, the perception of a high crime rate was associated with lower MVPA levels (β = −23.383, p = 0.002). Reporting the presence of zebra crossings and pedestrian signals or attractive natural sightings was associated with higher MVPA levels; however, this association varied across sociodemographic groups. These findings emphasize the importance of addressing air pollution and improving the built environment to facilitate outdoor active living, including active transportation – solutions that are particularly relevant not only for NCD risk mitigation, but also for climate change adaptation.

## INTRODUCTION

The health benefits of physical activity have been extensively researched, with studies associating physical activity with improved physical, psychological, social, and cognitive health indicators among children and youth (Castelli, 2022; Pascoe et al., 2020; Poitras et al., 2016). However, we are in the midst of a global physical inactivity pandemic (Kohl et al., 2012; Pratt et al., 2020) with recent studies indicating that a large proportion of children and youth do not meet the recommended levels of physical activity necessary to maintain good health (Aubert et al., 2022; Author, 2018; Author et al., 2023; Kuzik et al., 2023). Physical inactivity has not only been linked to the growing prevalence of chronic diseases such as heart disease, type-2 diabetes, and cancer (CDC, 2022), but it is also considered to be the fourth leading risk factor for mortality (WHO, 2012).

This is a pressing concern, as the burden of disability and mortality of non-communicable diseases (NCDs) has risen among children and youth (Akseer et al., 2020), with physical inactivity as a key risk factor (Budreviciute et al., 2020). Low to middle-income countries such as India experience a significant burden of NCDs, which is compounded by the presence of multiple syndemics (Frumkin and Haines, 2019; World Health Organization, 2009). These syndemics result from the coexistence and interaction of infectious diseases and NCDs, which further intensify health challenges in low to middle-income countries (Boutayeb, 2006; Oni and Unwin, 2015). Socioeconomic factors, including poverty, limited healthcare access, and inadequate infrastructure in many low to middle-income countries also exacerbate the burden of NCDs and infectious diseases (Gouda et al., 2019). In India, NCDs are a major health concern, accounting for nearly 60% of all deaths (Nethan et al., 2017).

Studies have found that behaviours such as physical activity can be effective in preventing NCDs (Budreviciute et al., 2020), thus promoting physical activity can not only help prevent a substantial number of NCD-related premature deaths, but also the overall burden on healthcare systems and society. In particular, meeting the globally recommended 60 minutes of daily moderate-to-vigorous physical activity (MVPA) is an effective strategy for preventing and managing NCDs (Baran et al., 2020; Sprengeler et al., 2021). A recent study indicated that a one-hour-per-day increase in light-intensity physical activity resulted in 10% less mortality, while the same one-hour-per-day increase in MVPA resulted in 40% less mortality among adults over the age of 40 (SaintLJMaurice et al., 2018). As a result, identifying the socioeconomic and environmental factors that influence physical activity levels, particularly MVPA, is imperative to help create conducive environments which reduce the NCD burden over the lifespan.

Air pollution is one environmental issue that influences outdoor physical activity, particularly in urban centres of India (Singh et al., 2021). Due to its recognized impact on physical and respiratory health, high levels of air pollution can deter outdoor activity among children and youth (Yu and Zhang, 2023). Multiple studies have found air pollution to be associated with increased odds of physical inactivity among children and adults (An et al., 2018), with one study identifying air pollution to be associated with lower physical activity and higher sedentary behaviour among children and youth (Yu and Zhang, 2023).

In addition to air pollution, the built environment can influence the health behaviours of children and youth. Built environment factors such as poor pedestrian safety, traffic hazards, and prevalence of crime have been associated with lower physical activity levels among children and youth in low-and middle-income countries (Hermosillo-Gallardo et al., 2020; Lv and Wang, 2023; Odunitan-Wayas et al., 2021), as well as in high-income countries such as Canada (Hunter et al., 2022). In contrast, built environment factors such as neighbourhood aesthetics have been associated with higher levels of physical activity among children and youth (Lv and Wang, 2023; Odunitan-Wayas et al., 2021; Qu et al., 2021).

Despite evidence indicating the growing impact of air pollution and the built environment on physical activity, there is a lack of research in India capturing the perceptions of children and youth regarding air pollution and the built environment in their communities. While other studies have focused specifically on the impacts of air quality or built environment features on MVPA (Author et al., 2023; Rees-Punia et al., 2018; Singh et al., 2021), this is the only study to date exploring location-specific experiences of air pollution, built environment, and MVPA from a youth perspective. Irrespective of location, most physical activity studies have focused on the perspectives of parents or caregivers, overlooking the unique viewpoints and concerns of younger populations (Daniel et al., 2020; Fan et al., 2017; Hunter et al., 2022; Wang et al., 2022; Waters et al., 2021). Understanding how children and youth perceive their environment is crucial in designing effective interventions and strategies to promote physical activity. More importantly, in the most populous country in the world, with diverse urban and rural environments of varying scales (Hertog and Gerland, 2023; Sciubba, 2023), it is important to understand the perception of children and youth across jurisdictions. The objective of this study is to understand the relationship between child and youth perceptions of air pollution, built environment, and MVPA across rural and urban regions in India.

## METHODS

### Design

A cross-sectional, observational study was conducted during the Coronavirus disease lockdown in India in 2021. Data were collected through online surveys as a part of a multi-center cohort study (Author et al., 2022; Vispute et al., 2023). The study implemented a multi-stage stratified random sampling method involving urban and rural schools in five Indian states across 28 different cities and villages as shown in **Figure 1**. Ethics approval was obtained from the Ethics Committee Jehangir Clinical Development Centre Pvt. Ltd in Pune, Maharashtra (EC registration number –ECR/352/Inst/MH/2013/RR-19). All study methods adhered to the Declaration of Helsinki for biomedical research involving human participants.

**Figure 1.**
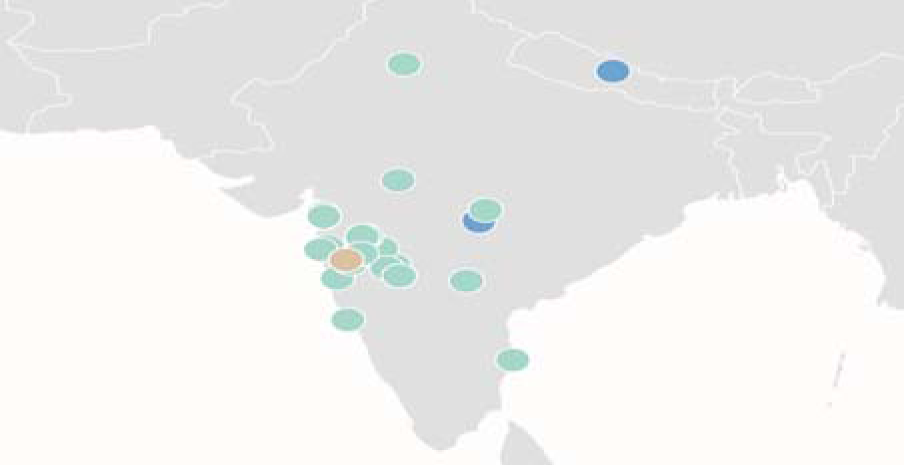
– Map of India showing 28 cities and villages sampled in the study.

### Recruitment and Participants

School principals (n = 50) from urban and rural areas across five Indian states (Maharashtra, Gujarat, Telangana, Madhya Pradesh, and Tamil Nadu) were invited to participate in the study. Of the invited principals, 41 agreed to participate and electronically shared the study information and consent forms with parents and children from their respective schools. Informed consent was obtained from 1042 children and youth aged 5 to 17 years and their parents. After receiving consent, anonymized questionnaires were administered via Google Forms between March 15, 2021, and May 20, 2021. Children under 13 years of age received parental assistance in completing the questionnaire. Children and youth were asked to self-report information on various factors, including physical activity and sedentary behaviour, perception of air pollution, participation in sports, active transportation, peer support, built environment, and school policies promoting physical activity.

### Sociodemographic Characteristics

In order to assess the relationship between various sociodemographic characteristics and behaviours, this study collected information on participants’ age, gender, and geographic location. Gender was collected by asking participants to report whether they are male or female. Children and youth provided their date of birth, which was used to calculate the age of each participant at the time of the study. As schools from both urban and rural centres were selected from five Indian states, the urban or rural residence was determined based on the location of the respective school in a city or village.

### Physical Activity Measures

To assess physical activity levels, participants were asked to indicate the frequency of their involvement in various activities. Children and youth completed a questionnaire with an extensive list of activities, including sports, weight training, running, and a range of other activities like jumping rope, playing garden games, and swimming. For each selected activity, participants reported the duration in minutes spent per day. The collected physical activity data were then categorized into three intensity types: inactivity, light activity, and MVPA [28,29]. In this study, MVPA was calculated by summing the daily average time spent in moderate and vigorous activities, including sports, garden games, running, aerobic activity, cycling, swimming, and weight training.

### Air Pollution Measures

Participants were asked to indicate their level of agreement using a 5-point Likert scale (strongly disagree, somewhat disagree, neither agree nor disagree, somewhat agree, strongly agree) with four statements related to air pollution in their neighbourhood. These statements included whether there is an air pollution problem, whether air pollution prevents them from being active outside, whether their parents restrict their outdoor activity due to air pollution, and if they take precautions against air pollution when going outside or engaging in outdoor physical activity. Responses obtained in this study using a 5-point Likert scale were analyzed as dichotomous variables. Responses falling within the “agree” category included “strongly agree” and “agree”, while responses falling within the “disagree” category included “strongly disagree” and “disagree.” If responses were “neither agree nor disagree,” they were excluded from the analyses.

### Built Environment Measures

Perceptions about the built environment were collected by asking children and youth about their level of agreement using a 5-point Likert scale (strongly disagree, somewhat disagree, neither agree nor disagree, somewhat agree, strongly agree) with statements such as: “There are attractive natural sightings in my neighbourhood,” “There is a high crime rate in my neighbourhood,” and “There are zebra crossing and pedestrian signals to help walkers cross busy streets in my neighbourhood.” The full list of questions can be found in the **Supplementary File**.

### Peer Support Measures

To assess peer support, participants were asked about the physical activity levels of their closest friends: "Your closest friends are the ones you enjoy spending the most time with. How many of your closest friends engage in regular physical activity?" Participants were provided with response options ranging from zero to four or more friends.

### Statistical Analysis

All statistical analyses were performed using R Studio version 2022.12.0+353 (RStudio Team, 2015). The primary independent variable was the perception of air pollution as a problem in the city, while the primary dependent variable was the average daily minutes spent on MVPA. Following established procedures for analyzing physical activity data, participants reporting over two standard deviations of average daily MVPA were considered outliers and removed from the analysis. The sample was divided based on gender (male and female), geographic location (urban and rural), and age cohorts (5-10 years old, 11-13 years old, and 14-17 years old), resulting in eight multiple linear regression models, including the overall model that used data from all participants. The estimated coefficients in these models indicated the expected change in daily MVPA associated with the participant’s perception of air pollution as a problem while controlling for age, community and built environment, and peer support. The gender models were also adjusted for location, while the location models were adjusted for gender. T-tests for unequal variances were conducted to compare average MVPA across various demographic and behavioural factors. A chi-square analysis was also conducted to compare the frequency distribution of responses to the perception of air pollution as a problem. All reported results in this study were considered statistically significant at a threshold of p < 0.05.

## RESULTS

The study consisted of 1042 participants. A total of 50 participants were excluded as their age was above the study’s target range of 5-17 years. The overall sample was further segregated by sociodemographic characteristics to assess differences in perceptions of air pollution across subgroups. As shown in **Table 1**, the study included 499 males (49.7%) and 493 females (50.3%), comprising 592 urban residents (59.7%) and 400 rural residents (40.3%). There were 339 participants aged 5 to 10 years (34.2%), 318 participants aged 11 to 13 years (32.1%), and 335 participants aged 14 to 17 years (33.8%).

**Table 1.** Children and youth sample summary by gender, age, and location (N = 992)

|  | **Total (N=992)** |  |
| --- | --- | --- |
| **Sample characteristic** | **N** | **%** |
| **Age Group** |  |  |
| 5 to 10 years | 339 | 34.2 |
| 11 to 13 years | 318 | 32.1 |
| 14 to 17 years | 335 | 33.8 |
| **Gender** |  |  |
| Male | 499 | 49.7 |
| Female | 493 | 50.3 |
| **Location** |  |  |
| Rural | 400 | 40.3 |
| Urban | 592 | 59.7 |

Of the total participants, 663 were included in the regression models after removing missing data and outliers for MVPA. As shown in **Table 2**, the average daily MVPA for all children and youth was 82.5 minutes. Those who perceived air pollution as a problem had lower MVPA levels compared to those who did not perceive air pollution as a problem (t (757.22) = −2.306, p = 0.033). Males had significantly higher MVPA levels compared to females (t (917.92) = 10.571, p < 0.001), There was no significant difference in MVPA levels between urban and rural areas; however, more urban residents reported an air pollution problem than rural residents (χ² = 87.297, p < 0.001). Moreover, among participants from rural areas, those who perceived an air pollution problem had lower MVPA levels compared to those who disagreed (t (355.36) = 2.3762, p = 0.018). When examining MVPA levels across different age groups, there was no significant difference observed.

**Table 2.**
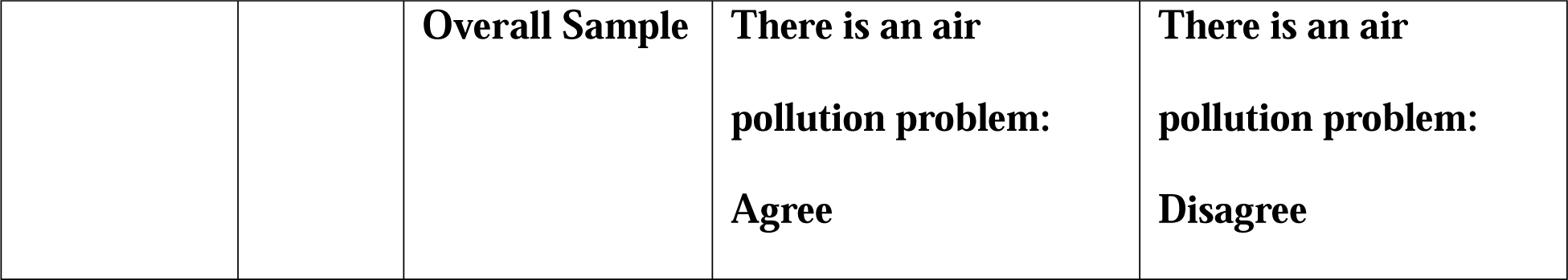
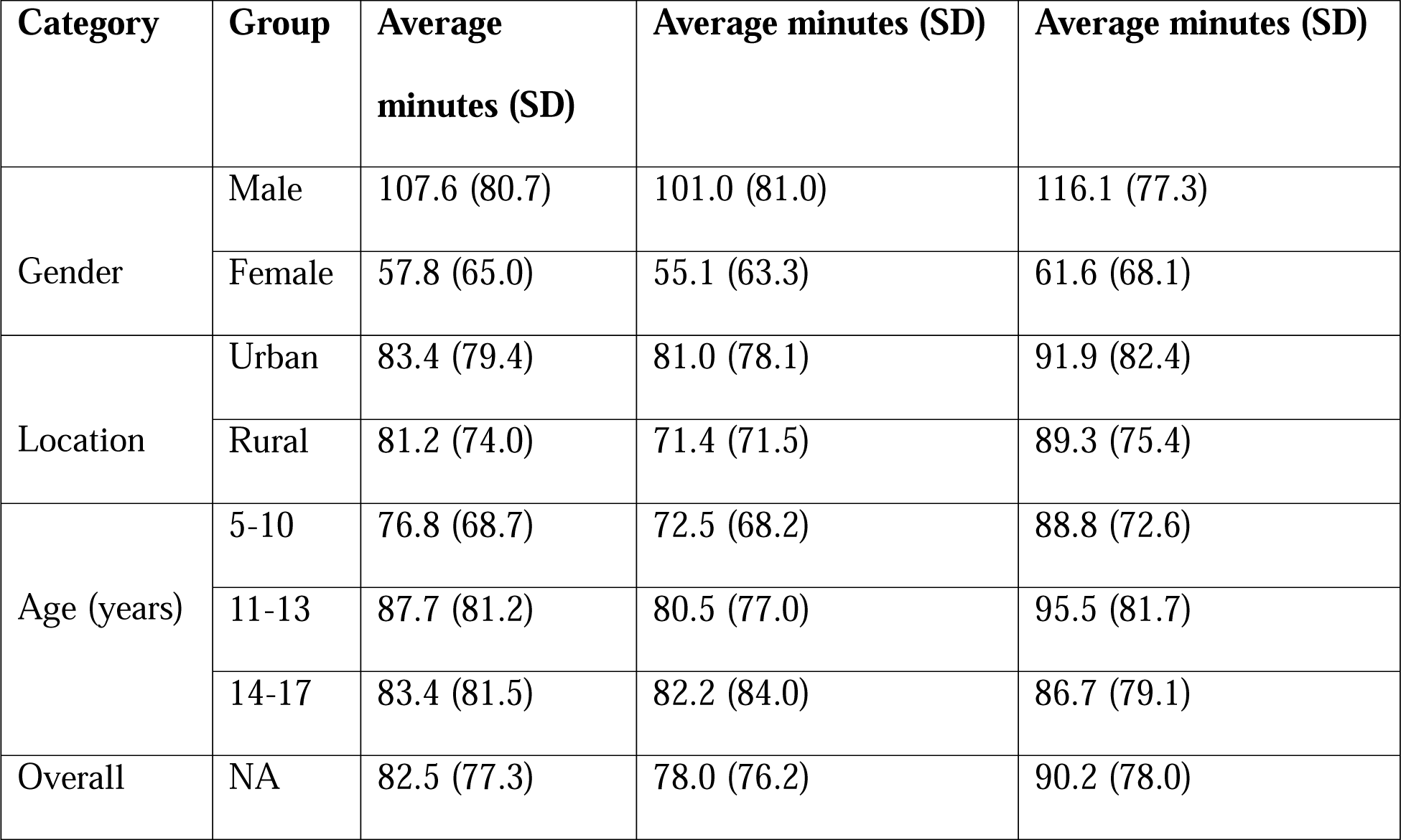
Daily minutes of engagement in MVPA among children and youth based on gender, age, and location.

**Table 3** shows the association between air pollution and MVPA for the full sample of children and youth. Participants who agreed that there is an air pollution problem were associated with lower MVPA levels (β = −18.365, p = 0.002), compared to those who disagreed. Furthermore, the presence of a high crime rate in the neighbourhood was also associated with lower MVPA levels (β = −23.383, p = 0.002) compared to those who disagreed. Having one or more active friends was associated with higher levels of MVPA (β = 33.531, p = 0.005) in comparison to having no active friends. However, other built-environment factors such as the presence of footpaths, attractive natural sightings, zebra crossings, and pedestrian signals did not show significant associations with MVPA levels in the overall model.

**Table 3.** Relationship between reporting that there is an air pollution problem and moderate to vigorous physical activity (Model 1)

|  | **Beta coefficients (95% Confidence Intervals)<sup>a</sup>** |
| --- | --- |
| There is an air pollution problem: Agree | -18.365* (-30.115, -6.615) |
| **Disagree – (Ref)** |  |
| There is a high crime rate in my neighborhood: Agree | -23.383* (-37.856, -8.910) |
| **Disagree – (Ref)** |  |
| There are attractive natural sightings in my neighborhood: Agree | -1.532 (-13.623, 10.558) |
| **Disagree – (Ref)** |  |
| There are zebra crossings and pedestrian signals to help walkers cross busy streets in my neighborhood: Agree | 10.590 (-3.038, 24.219) |
| **Disagree – (Ref)** |  |
| Has one or more active friends: Yes | 33.531* (10.354, 56.708) |
| **Has no active friends – (Ref)** |  |
| Constant | 7.980 (-25.378, 41.338) |

\* Indicates a statistically significant relationship at the $p < 0.05$ level
<sup>a</sup>The model was adjusted for age, gender, and location.

**Table 4** demonstrates that across the three age groups, significant associations were found between reporting air pollution and MVPA levels. In the 5 to 10-year age group, the perception of an air pollution problem was associated with lower levels of MVPA (β = −26.725, p = 0.012)

When comparing the models by gender, in the male model, agreeing that air pollution is a problem was associated with higher MVPA levels (Males: β = −25.090, p = 0.008); however, this association was not significant in the female model. Among females, the presence of a high crime rate in their neighbourhood was associated with lower MPVA levels (Females: β = - 27.620, p = 0.001), however; this relationship was not significant for males. In contrast, having one more active friend was associated with higher MVPA levels in females (β = 40.085, p = 0.001).

**Table 4.**
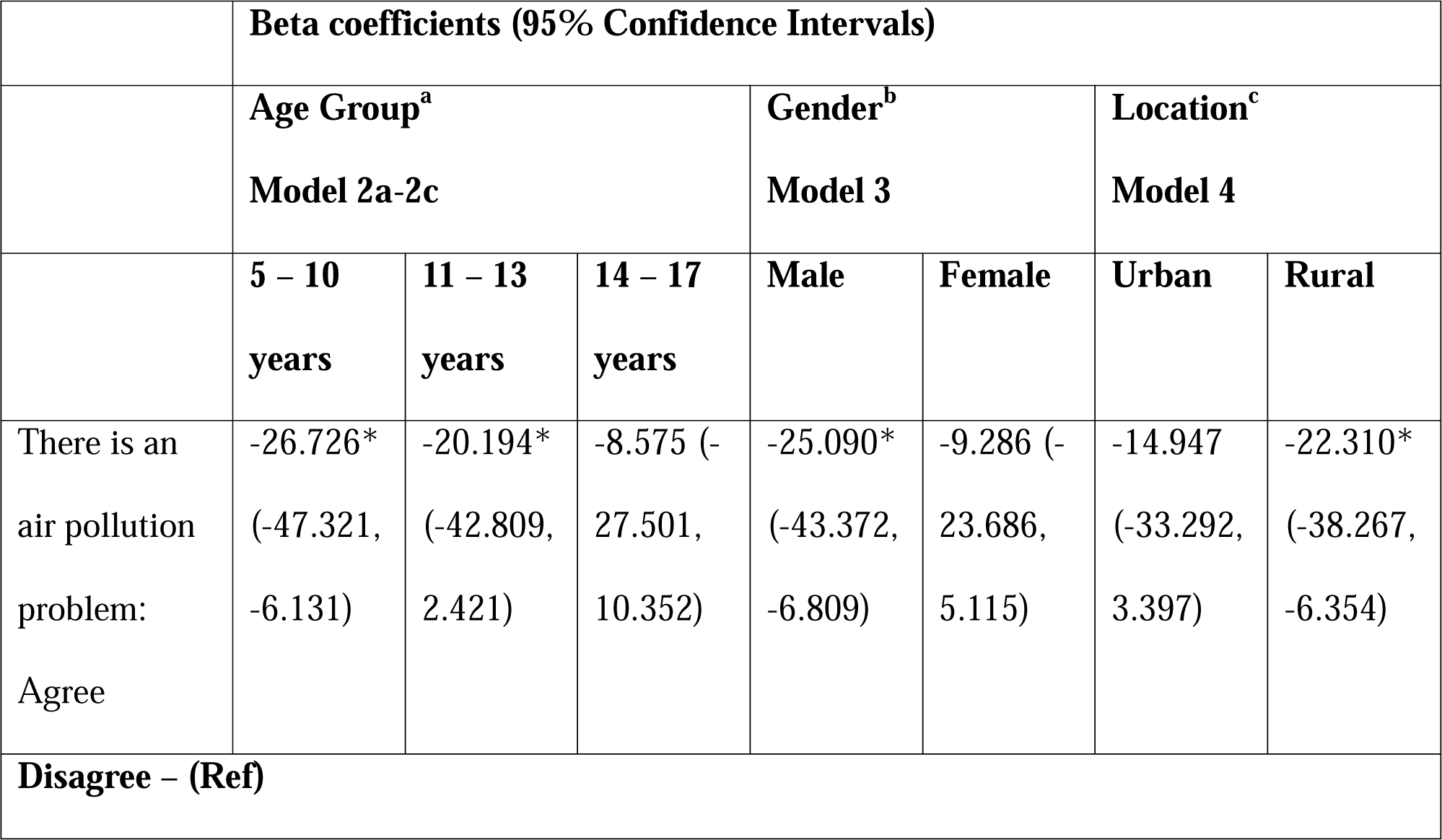
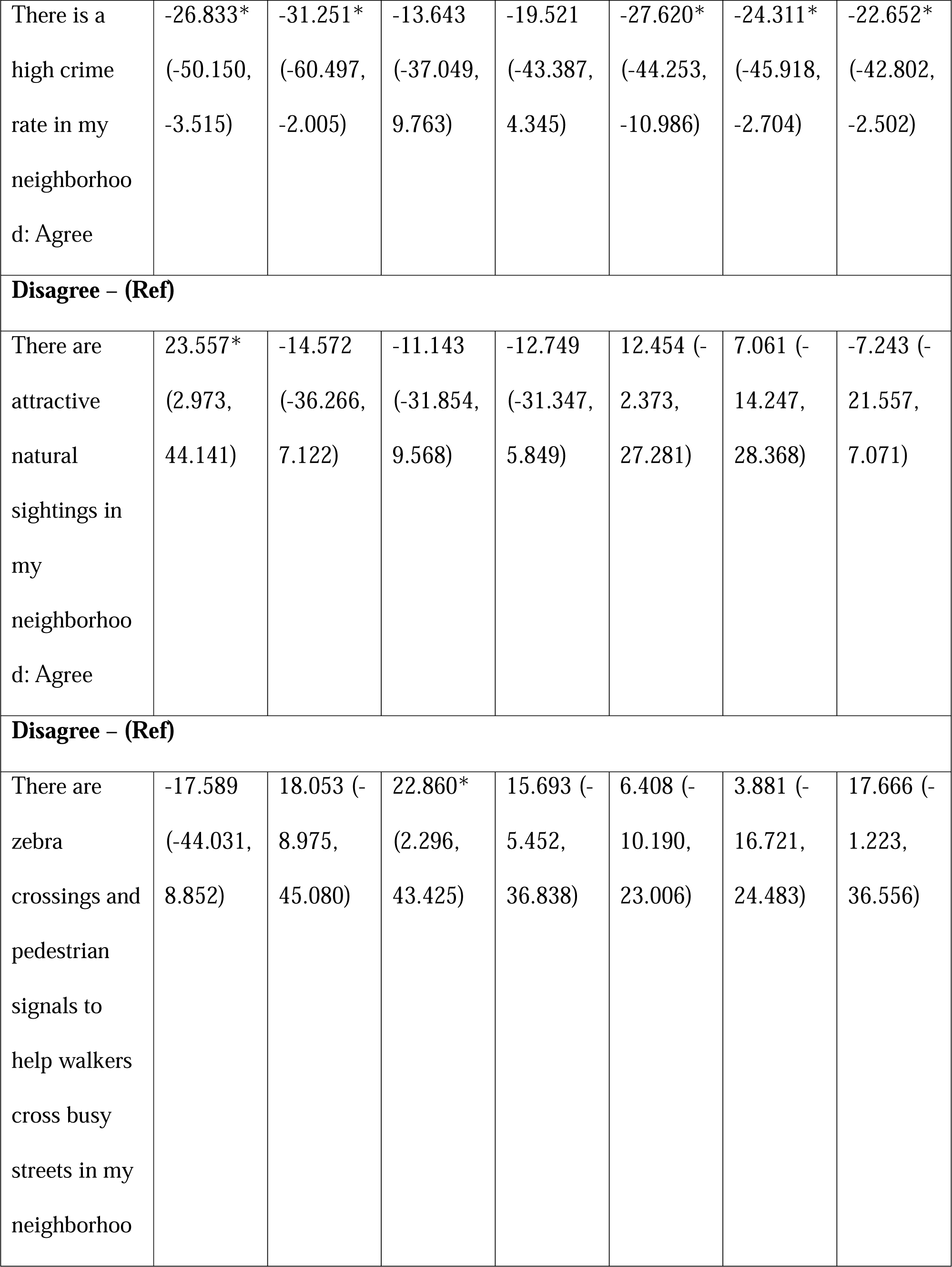
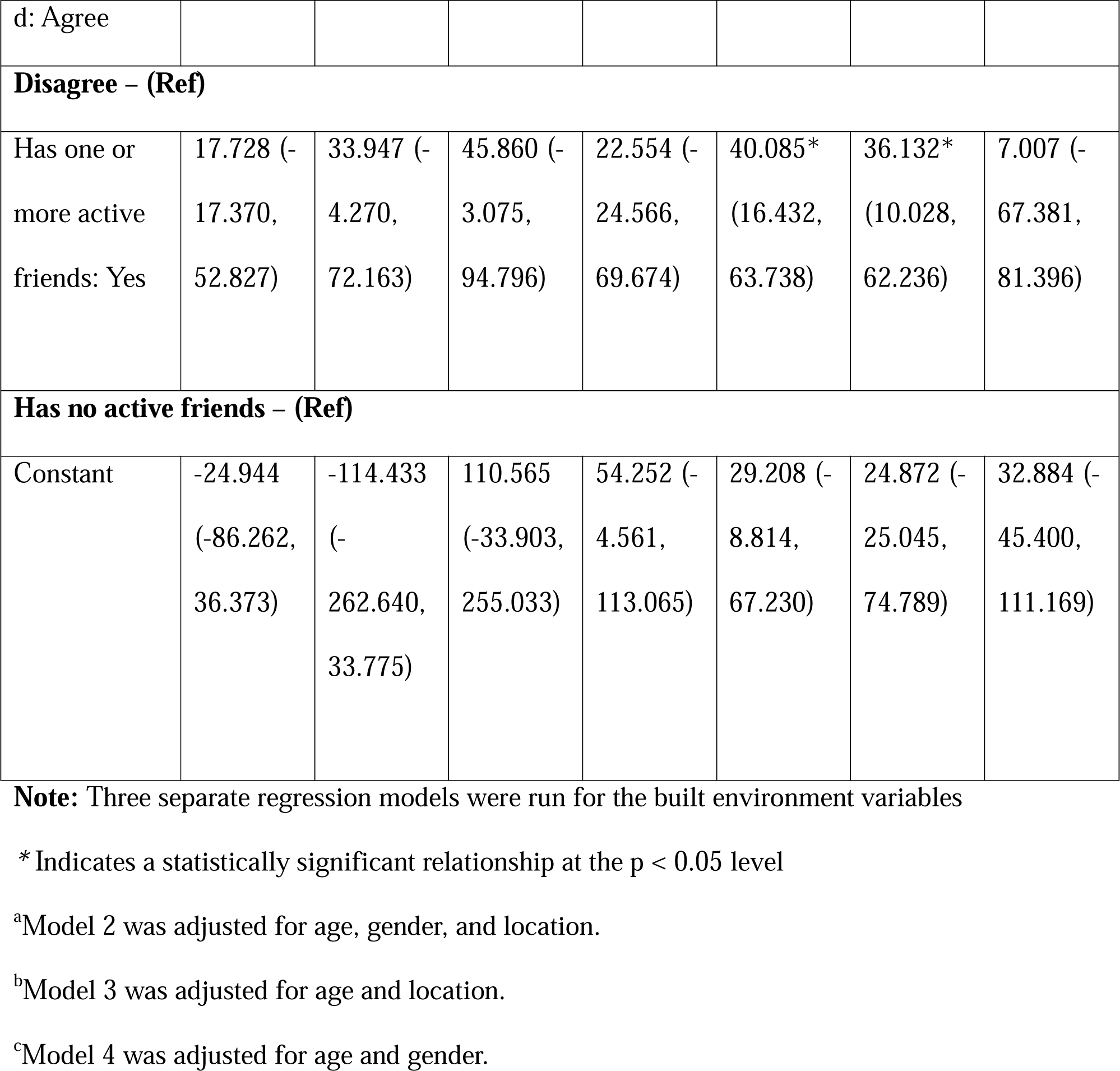
Relationship between reporting that there is an air pollution problem and moderate to vigorous physical activity across various sociodemographic groups.

In the urban model, agreeing with the statement that air pollution is a problem was not significantly associated with MVPA levels. However, in the rural model, the perception of air pollution as a problem was associated with lower MVPA levels (β = −22.310, p = 0.006), compared to those who did not perceive air pollution to be a problem. Contrastingly, having one or more active friends was associated with more MVPA in the urban model (β = 36.132, p = 0.007) but not in the rural model. Further, reporting high crime was associated with lower MVPA levels in both the rural model (β = −22.652, p = 0.028) and the urban model (β = −24.311, p = 0.028).

## DISCUSSION

This is the first study to investigate the relationship between child and youth perceptions of air pollution, built environment, and MVPA in urban and rural regions in India. In building on the key findings and gaps identified in the 2022 India Report Card on Physical Activity for Children and Adolescents (Author et al., 2023), this study focused on perceptions of children and youth that have thus far been largely ignored in active living research in India.

Given the wide-ranging impacts of air pollution on vulnerable groups such as children and youth, there is an urgent need to understand how this group’s physical activity levels are affected by air pollution, especially considering the worsening air quality in India that is being attributed to climate change (Singh et al., 2023; Van Oldenborgh et al., 2018). By including the perceptions of children and youth, this study provides a distinct understanding of the impact of air pollution and the built environment on physical activity in younger populations.

The overall findings suggest that perceptions of an air pollution as a problem and negative perceptions of the built environment are both associated with lower levels of MVPA among children, a finding that aligns with active living research in other parts of the world (Goon et al., 2020; Gu et al., 2023; Loh et al., 2019; Taylor et al., 2018). This finding suggests that while air pollution is generally high in Indian cities and towns (Dandotiya et al., 2020; Hussain et al., 2023; Mahendra et al., 2023), perception of air pollution -- irrespective of actual levels of air pollution -- can potentially influence the physical activity of children and youth.

As the evidence indicates that there are differences in not only air pollution, but also MVPA across urban and rural areas in India (Author et al., 2023; Vieira et al., 2023), we developed statistical models segregated by location (urban vs. rural). More children and youth in urban areas perceived air pollution as a problem, a finding that aligns not only with current evidence of air pollution perceptions (Author et al., 2023), but also with actual air pollution levels in India that tend to be higher in urban areas (WHO, 2023). However, contrastingly, rural children and youth who perceived higher air pollution reported significantly lower MVPA in comparison with children and youth who reported lower perception of air pollution – a finding that was not significant in urban areas. This finding also contrasts with previous research which indicates that rural residents lack awareness of the impacts of air pollution on health (Yang, 2020). It is plausible that children and youth in urban areas may get habituated or desensitized to the level of pollution and stop perceiving it as problematic. On the other hand, it is also possible that pollution levels are increasing in rural areas of India due to rapid industrialization (Karambelas et al., 2018; Ravishankara et al., 2020). These findings require further exploration, including linking citizen perception data with objective weather data.

In addition to location-based differences, by examining perceptions of air pollution and MVPA levels in males and females, this study adds to the existing evidence on gender-based differences in physical activity engagement (Author, 2018; Author et al., 2023). Males reported higher MVPA compared to females, a finding that aligns with existing literature that suggests males in India tend to accumulate higher levels of physical activity compared to females, driven by factors such as social acceptability, cultural norms, and general perception of lack of safety that limit outdoor activities among female children and youth neighbourhoods (Author et al., 2023; Mathur et al., 2021; Tyagi and Raheja, 2020). Among males, the perception of air pollution as a problem was associated with lower levels of MVPA; however, this association was not significant in females, a finding that further suggests that female children and youth might not be obtaining as much MVPA outdoors in comparison to males. These findings indicate the need for targeted interventions to promote physical activity in both male and female populations.

The study findings also reveal significant associations between the perception of air pollution and MVPA levels among children and youth in different age groups. Perceiving air pollution to be a problem was linked to lower MVPA in the 5 to 10-year age group, but not in the 11 to 13 and 14 to 17-year age groups. These findings could be due to younger children being more influenced by their immediate surroundings and parental guidance, and likely having less independent mobility than older children (Pickhardt, 2010; Schoeppe et al., 2013). Moreover, studies have shown younger children to be more physiologically susceptible to the negative effects of air pollution, which may lead to changes in their physical activity behaviours (Landrigan et al., 2010; Sunyer et al., 2015; UN, 2018).

In terms of the built environment, the presence of attractive natural sightings in the neighbourhood was related to higher MVPA in the 5 to 10-year age group. One potential explanation for this association is young children’s fascination with novel sightings in nature, as engaging with nature supports imaginative play and interaction with their environment (Dowdell et al., 2011). Similarly, community-related factors such as perception of high crime rates in the neighbourhood were associated with reduced MVPA in the 5 to 10 and 11 to 13-year age groups, but not among 14 to 17-year-olds. It is possible that older adolescents might develop different coping mechanisms for dealing with perceived environmental challenges and have learned to adapt to their surroundings when making decisions about physical activity engagement (Dwyer et al., 2012). Furthermore, among females, the significant association between a high crime rate in their neighbourhood and lower MVPA levels highlights the potential impact of environmental safety concerns on physical activity. This finding is consistent with research indicating that safety concerns, particularly for women, can act as a barrier to physical activity, limiting opportunities for outdoor exercise and leading to reduced overall activity levels (Author et al., 2023; Mathur et al., 2021; Tyagi and Raheja, 2020). This discrepancy emphasizes the need for gender-specific interventions and strategies to address safety concerns and promote physical activity among females.

In this study, the presence of zebra crossings and pedestrian signals was associated with increased levels of MVPA among children and youth aged 14 to 17; however, this association was not observed in the younger population. These findings could potentially be explained by older adolescents having more independent mobility, including their choice of transportation and routes. As zebra crossings provide safer and more convenient opportunities for pedestrians to cross busy streets (Ismail et al., 2023), they may encourage older adolescents to engage in more outdoor physical activity. In contrast, younger children might rely more on their parents or guardians to dictate their travel routes (Pickhardt, 2010; Schoeppe et al., 2013), and the presence of zebra crossings may not have the same impact on their physical activity as with older adolescents. These findings demonstrate the need for policymakers to prioritize the implementation and maintenance of zebra crossings and pedestrian signals as a means to promote physical activity, particularly in older adolescents.

Furthermore, the impact of peer support on MVPA was found to vary based on gender and location. Previous research has found that by facilitating social connections and promoting enjoyable physical activities, active friends play a vital role in enhancing both physical and mental well-being (Author et al., 2023; Jago et al., 2011; Leggett et al., 2012). In this study, female and urban children and youth who reported having active friends had higher MVPA; however, the same association was not observed in males or rural residents, indicating that other factors might influence their MVPA levels, i.e., the impact of peer support on their physical activity behaviours may be different from those observed in females or urban residents. Further research is required to better understand the specific mechanisms and contextual factors that contribute to these disparities among different demographic groups in India.

The study findings on perceptions of built environmental factors, perceptions of air pollution, and demographic-related variations in physical activity are particularly relevant in light of climate change impacts, with rising temperatures and heat waves exacerbating the effects of air pollution, particularly in the global south (Singh et al., 2023; Van Oldenborgh et al., 2018; Yang and Shao, 2021). In low to middle-income countries such as India where there are increasing levels of particulate matter pollution due to industrial emissions, vehicle exhaust, and the burning of biomass, understanding how people perceive and respond to air pollution is not only crucial for promoting health in children and youth, but is also important for NCD prevention and management (Gargava and Rajagopalan, 2016; India State-Level Disease Burden Initiative Air Pollution Collaborators, 2019; Upadhyay et al., 2018). Nevertheless, with the impact of climate change being felt across the world (Dahl et al., 2023; Hanes et al., 2019), irrespective of location, child and youth perceptions are critical in understanding changing patterns of MVPA globally – an area of research that will require more attention going forward.

### Strengths and Limitations

By examining the unique perceptions of air pollution among children and youth across 28 cities and villages from urban and rural populations, this study allowed for cross-jurisdictional comparisons. However, as data were collected at one time point during the COVID-19 pandemic lockdown, reported pollution perceptions and physical activity may differ from a non-pandemic period. Future studies should focus on conducting longitudinal data collection, where both objective air pollution and MVPA data should be linked with the subjective perception of children and youth.

## CONCLUSION

This is the first study to investigate the association between child and youth perceptions of air pollution, built environment, and MVPA in both urban and rural regions of India. The study findings emphasize the importance of considering children and youth’s unique perceptions of air pollution and their built environment, as they significantly impact their physical activity levels. The study’s findings not only highlight the location-, gender-, and age-specific criteria in informing targeted physical activity interventions, but the results indicate the need to focus on child and youth perceptions of air pollution in this age of climate emergency, irrespective of location to understanding changing patterns of MVPA.

## Data Availability

All data produced are available online at doi.org/10.6084/m9.figshare.23993022

