## Supplemental Table 1 for "The interplay between air pollution, built environment, and physical activity: perceptions of children and youth in rural and urban India"

**Physical Activity Questionnaire**

1. Average minutes spent in toilet (dressing, undressing, and showering):
   a. 15
   b. 30
   c. 45
   d. 60+
2. Average minutes spent in making bed:
   a. 15
   b. 30
   c. 45
   d. 60+
3. Average minutes spent having breakfast, lunch, dinner, and any other meal:
   a. 15
   b. 30
   c. 45
   d. 60+
4. Average minutes spent studying or doing homework at home/tuition daily:
   a. 5
   b. 30
   c. 45
   d. 60
   e. 75
   f. 90
   g. 120+

**Which other activities do you do at school/ after school?**

| Section Name | Activity Name | Duration in actual minutes | Frequency/ week  1 / 2 / 3 / 4 / 5 / 6 / 7 |
| --- | --- | --- | --- |
| Inactivity | Chatting with friends |  |  |
|  | Watching television |  |  |
|  | Internet surfing (Facebook, Instagram, Youtube, reading news, etc.) on a desktop/laptop/tablet |  |  |
|  | Internet surfing (e.g. Facebook, Snapchat, Instagram, Youtube, Tiktok, reading news, etc.) on a smartphone.  *This does NOT include regular texting or gaming time.* |  |  |
|  | Playing games on a desktop/laptop/gaming device/television |  |  |
|  | Playing video games on a smartphone |  |  |
|  | Playing video games on a handheld video game console |  |  |
|  | Work on desktop/laptop/tablet |  |  |
|  | Texting/whatsapp using a smartphone |  |  |
| Light Activity | Daily household work |  |  |
|  | Reading |  |  |
|  | Doll games |  |  |
|  | Playing board games/Sitting games |  |  |
|  | Breathing yoga (eg. Pranayama) |  |  |
|  | Light walking |  |  |
|  | Mopping |  |  |
|  | Washing clothes |  |  |
|  | Cooking |  |  |
|  | Taking care of siblings |  |  |
| Moderate activity | Running games |  |  |
|  | Garden games (slides/jungle gym) |  |  |
|  | Cycling |  |  |
|  | Swimming |  |  |
|  | Jump ropes |  |  |
|  | Brisk walking |  |  |
|  | Yoga (physical practices such as Ashtanga or Vinyasa) |  |  |
|  | Collecting water from well |  |  |
|  | Plucking tea leaves |  |  |
| Vigorous activity | Karate (Martial Arts) |  |  |
|  | Badminton |  |  |
|  | Football |  |  |
|  | Tennis |  |  |
|  | Baseball |  |  |
|  | Volleyball |  |  |
|  | Basketball |  |  |
|  | Aerobics |  |  |
|  | Weight training (e.g., push-ups, sit-ups) |  |  |
|  | Kho kho |  |  |
|  | Kabbaddi |  |  |
|  | Skating |  |  |
|  | Cricket |  |  |
|  | Horse Riding |  |  |
|  | Running |  |  |
|  | Golf /Squash |  |  |
|  | Dancing specify - |  |  |

**School:**

1. Does your school organize physical activities before-school hours, during lunch hour, or after-school hours?
   a. Yes
   b. No
   c. If yes, do you participate in them? _________________________
2. Can you use INDOOR physical activity areas at school between classes? (e.g., using dance studio or yoga space during lunch, spare periods)
   a. Yes
   b. No
   c. I don't know
3. Can you use OUTDOOR physical activity areas at school between classes? (e.g., using playing field during lunch, spare periods)
   a. Yes
   b. No
   c. I don't know
4. Does your school have:
   a. Gymnasium(s) - Yes / No
   b. Indoor facilities (e.g., dance studio, yoga room, fitness room) - Yes / No
   c. Outdoor facilities (e.g., playing fields, paved activity areas) - Yes / No
5. Can you use physical activity equipment throughout the school day? (e.g., footballs, badminton racquets)
   a. Yes
   b. No
   c. I don't know
6. Before or after school, can you use the following spaces at your school? (i.e., early mornings, evenings, weekends)
   a. Gymnasium(s) - Yes / No
   b. Indoor facilities (e.g., dance studio, yoga room, fitness room) - Yes / No
   c. Outdoor facilities (e.g., playing fields, paved activity areas) - Yes / No
   d. Equipment (e.g., Footballs, basketballs) - Yes / No
7. Does your school have sports/physical activity competitions with other schools? (E.g. Cricket, hockey, football)
   a. Yes
   b. No
   c. If yes, do you participate? _________________________

**Family and Peers:**

1. Who do you play sports or do other physical activities with? Please select all that apply.
   a. Parents
   b. Siblings
   c. Cousins/other family
   d. Neighbours
   e. Peers/friends from school
   f. Peers/friends from outside of school
   g. Other (please specify): _______________________
2. How much do your parents or guardians encourage you to be physically active?
   a. Strongly encourage
   b. Sometimes encourage
   c. Do not encourage
3. How much do your parents or guardians support you in being physically active? (e.g., taking you to team games, buying you sporting equipment)
   a. Strongly support
   b. Sometimes support
   c. Do not support
4. Your closest friends are the friends you like to spend the most time with. How many of your closest friends are physically active?
   a. None
   b. 1 friend
   c. 2 friends
   d. 3 friends
   e. 4 friends

**Perception of Environment:**

Please circle the answer that best applies to you and your neighborhood.

1. There is an air pollution problem in my city.
   a. Strongly disagree
   b. Somewhat disagree
   c. Neither agree nor disagree
   d. Somewhat agree
   e. Strongly agree
2. Air pollution prevents me from being active outside.
   a. Strongly disagree
   b. Somewhat disagree
   c. Neither agree nor disagree
   d. Somewhat agree
   e. Strongly agree
3. My parents restrict my outdoor activity because of air pollution.
   a. Strongly disagree
   b. Somewhat disagree
   c. Neither agree nor disagree
   d. Somewhat agree
   e. Strongly agree
4. I take precautions against air pollution when I go outside/engage in outdoor physical activity.
   a. Strongly disagree
   b. Somewhat disagree
   c. Neither agree nor disagree
   d. Somewhat agree
   e. Strongly agree
5. There are footpaths on most of the streets in my neighborhood.
   a. Strongly disagree
   b. Somewhat disagree
   c. Neither agree nor disagree
   d. Somewhat agree
   e. Strongly agree
6. My neighborhood streets are well lit at night.
   a. Strongly disagree
   b. Somewhat disagree
   c. Neither agree nor disagree
   d. Somewhat agree
   e. Strongly agree
7. There are zebra crossings and pedestrian signals to help walkers cross busy streets in my neighborhood.
   a. Strongly disagree
   b. Somewhat disagree
   c. Neither agree nor disagree
   d. Somewhat agree
   e. Strongly agree
8. There are trees along the streets in my neighborhood.
   a. Strongly disagree
   b. Somewhat disagree
   c. Neither agree nor disagree
   d. Somewhat agree
   e. Strongly agree
9. There are attractive natural sights in my neighborhood (e.g., trees, parks, ponds, rivers).
   a. Strongly disagree
   b. Somewhat disagree
   c. Neither agree nor disagree
   d. Somewhat agree
   e. Strongly agree
10. There is so much traffic that it makes it difficult or unpleasant to walk in my neighborhood.
    a. Strongly disagree
    b. Somewhat disagree
    c. Neither agree nor disagree
    d. Somewhat agree
    e. Strongly agree
11. There is a high crime rate in my neighborhood.
    a. Strongly disagree
    b. Somewhat disagree
    c. Neither agree nor disagree
    d. Somewhat agree
    e. Strongly agree
12. The crime rate in my neighborhood makes it unsafe to go on walks during the day.
    a. Strongly disagree
    b. Somewhat disagree
    c. Neither agree nor disagree
    d. Somewhat agree
    e. Strongly agree
13. The crime rate in my neighborhood makes it unsafe to go on walks during the night.
    a. Strongly disagree
    b. Somewhat disagree
    c. Neither agree nor disagree
    d. Somewhat agree
    e. Strongly agree
14. How far is your school from home?
    a. <2km
    b. <5km
    c. <10km
    d. 20km
    e. 20km+
15. Do you ride your bike to school?
    a. Yes (if yes, how often?)
       i. One day per week
       ii. 2-3 days per week
       iii. 4-5 days per week
       iv. 6-7 days per week
    b. No
16. A) Do you walk to school?
    a. Yes (if yes, how often?)
       i. One day per week
       ii. 2-3 days per week
       iii. 4-5 days per week
       iv. 6-7 days per week
    b. No
17. If you do not bike or walk to school, please select your mode of transportation from home to school:
    a. Parent's/family car
    b. Bus
    c. Auto rickshaw
    d. Taxi
    e. Scooter/motor bicycle
    f. Other (please specify): __________________________
